# Fear, Anxiety, Stress, and Depression of novel coronavirus (COVID-19) pandemic among patients and their healthcare workers

**DOI:** 10.1101/2021.05.28.21258006

**Authors:** Ashwin Parchani, K Vidhya, Prasan Kumar Panda, Vikram Singh Rawat, Yogesh Arvind Bahurupi, Deepjyoti Kalita, Harsh Kumar, Naveen Dr

**Author notes:** Emails in sequence. **Corresponding Author**: Prasan Kumar Panda, Asst. Professor, Dept. of Medicine (Infectious disease division), Sixth Floor, College Block, All India Institute of Medical Sciences (AIIMS), Rishikesh, India, 249203.

## Abstract

**Background:** Disease pandemics are known to cause psychological distress. The ensuing mental health issues are not only restricted to the patients and their relatives/friends but affect the healthcare workers (HCWs) as well. Our study aims to assess these psychological trends during the COVID-19 pandemic between the two most affected population groups, that is, patients and frontline healthcare workers.

**Methods:** A survey questionnaire including scales to assess fear, anxiety, stress, depression - PSS 10, and DASS 21 was distributed and sent to all COVID-19 suspected/confirmed individuals and healthcare workers at a tertiary care center along with a second visit after 14 days of answering the first questionnaire and this continued as follow up. Data were analyzed with the SPSS Version 23 using various tests of significance.

**Results:** In the community, COVID-19 patients in the age group 41-50 with respiratory tract symptoms and those who were home isolated/quarantined experienced a greater tendency of mental health problems. Healthcare workers posted in COVID-19 designated areas of the hospital displayed higher levels of stress, anxiety, and depression.

**Conclusion:** The high degree of uncertainty associated with novel pathogens has a profound effect on the psychological state of suspected/confirmed cases as well as healthcare workers. Within the community, individuals suspected of having COVID-19 display a significant mental health burden, while HCWs also experience an unprecedented amount of stress during such enduring situations.

**Key points:** *Question:* What is the psychological impact among patients and their healthcare workers during the COVID-19 pandemic?

*Findings:* In this observational study based on PSS 10 and DASS 21 questionnaire that included 156 patients and 226 health care workers, the patients in the age group 41-50 with respiratory tract symptoms and those who were home isolated/quarantined experienced a greater tendency of mental health problems. Similar burden was observed among health care workers.

*Meaning:* In a COVID-19 pandemic both population groups displayed higher levels of fear, anxiety, stress, and depression.

## Introduction

The novel coronavirus has spread across borders in a small-time affecting the world (1) and was announced to be a pandemic by WHO in March 2020 (2). The Coronavirus 19 initially appeared as a spurt of respiratory infections in Hubei city, China in December 2019. Widespread outbreaks are known to cause psychological distress and mental illnesses (3). Humanity has suffered in the hands of many such bio-disasters such as Severe Acute Respiratory Syndrome (SARS), Flu, Plague, Ebola, and Middle East Respiratory Syndrome (MERS), among others, with each of them promulgating a certain level of fear and mental stress among frontline workers and the affected population.

The effect on the community of the COVID-19 pandemic is also evident in studies. Ni et al. conducted a cross-sectional web-based study and found that among the 1577 community-based adults who participated in the study, about one-fifth of respondents reported probable anxiety (n=376, 23.84%, 95% CI 21.8-26.0) and probable depression (n=303, 19.21%, 95% CI 17.3-21.2). Within the community as well, patients afflicted with COVID-19 manifested higher levels of mental health burden. A gender effect was observed in the score of “Perceived Helplessness”, the subscale of PSS-10, with female patients showing higher scores compared to male patients, female, and male controls (4). Even among clinically stable COVID-19 patients, the prevalence of significant posttraumatic stress symptoms associated with COVID-19 was 96.2% (95% CI 94.8–97.6%) in a study (5). Nyuyen and colleagues assessed the level of depression in outpatients in Vietnam and interestingly witnessed that people with college/university or above educational attainment had higher odds of depression than those with elementary school/illiterate attainment. In addition, in comparison to people who had a low social status, were less healthy, had less physical activity, to those with middle or high social status, healthier, and had more physical activity had lower odds of depression (6).

These mental issues are not only restricted to patients and their close ones, but healthcare workers (HCWs) are also not immune to the stress caused by the spread of the disease (7). The high degree of uncertainty associated with novel pathogens has a profound effect on the psychological state of HCWs working at the forefront. HCWs experience an unprecedented amount of stress during such enduring situations. The extended and frequent changing shift times and risk of infection add to the pressure faced by these workers. The inability to effectively communicate while wearing personal protective equipment and poor performance of diagnostic procedures play a key role in the emergence of resentment in the minds of these brave warriors. A cross-sectional survey-based study during the COVID-19 pandemic illustrated that depression, anxiety, insomnia, and distress were widely prevalent among HCWs (8). Another study among HCWs (non-medical vs medical HCWs) revealed a higher prevalence with statistical significance of insomnia (38.4% vs. 30.5%), anxiety (13.0% vs. 8.5%), depression (12.2 vs. 9.5%), somatisation (1.6% vs. 0.4%), and obsessive-compulsive symptoms (5.3% vs. 2.2%) (9).

Our study emphasizes these psychological trends during the pandemic among the two most affected population groups, that is, the patient and contacts along with their treating frontline HCWs. Therefore, we aimed to study the psychological parameters of these two population groups in a tertiary care COVID-19 hospital.

## Objective

To assess the mental impact of 2019 novel coronavirus (Covid-19) outbreaks among COVID-19 patients (suspects and confirmed), their close contacts, and associated healthcare providers using a questionnaire-based study

## Materials and Methodology

### Settings and participants

The research was conducted at a tertiary care hospital in North India between March and November 2020. A structured questionnaire was distributed electronically to all participants. The study was conducted in compliance with the protocol and ethical approval was obtained from the institutional ethical committee (CTRI/2020/05/025494). Before taking the survey, the respondents signed a consent document stating that they had provided their informed written consent. There were no benefits, and participation was on an opt-in voluntary basis at the COVID-19 screening outpatient department. Data security and anonymity were ensured.

#### Inclusion criteria

- All COVID-19 suspected/confirmed patients and high-risk contacts (family and friends) reporting screening of OPD in a tertiary care hospital between March and November 2020
- All healthcare workers directly or indirectly related [treating or managing these patients]

#### Exclusion criteria

- Patients/subjects who did not consent
- Patients/subjects below the age of 18.

## Procedure/intervention

An online Google form was created for the questionnaire (Appendix 1 and 2) comprising PSS (Perceived Stress Scale) and DASS 21 (Depression Anxiety and Stress Scale) questionnaire. After procuring their phone number through hospital record within 24h of OPD visit, participants were contacted through WhatsApp or direct messaging or phone calls and instructed to properly read the consent form and questionnaire, after which they were provided the link for the form to be filled.

### PSS

The “PSS” suggested by Cohen et al. in 1983 measures the degree to which individual situations are evaluated as being stressful or more precisely, unpredictable, uncontrollable, and intense (overloading in 1983, then overwhelming in 1991). The tool is composed of 14 items on a five-point scale (going from “never” to “often”) (examples of items: “…have you ever been bothered by an unexpected event?” “… have you ever successfully coped with little problems and daily worries?”) (10).

### DASS 21

This was designed to measure emotional distress in three sub-categories (Lovibond and Lovibond, 1995) of depression (e.g. loss of self-esteem/incentives and depressed mood), anxiety (e.g. fear and anticipation of negative events), and stress (e.g. persistent state of over-arousal and low frustration tolerance). It is a self-reported questionnaire with 21 items (seven items for each category). To calculate comparable scores with full DASS, each 7-item scale was multiplied by two. Participants were asked to rate how many of each of the items (in the form of statements) applied to them over the past days, using a five-point scale (completely disagree to completely agree) (11).

### Comparator and Outcome

There was no comparator, and the outcomes were measures of fear, anxiety, stress, and depression among the participants.

### Statistical analysis

Data were analyzed using the latest version of SPSS software (Version 23). Fisher’s exact test and the chi-square test were used to correlate the association. The Stuart-Maxwell test was used to assess the change in response in the HCW population between the first point of contact and follow-up. Statistical significance was set at a P-value <0.05.

## Results

Of the 708 COVID-19 patients (suspects/confirmed/contacts) and 442 HCWs, 156 and 226 participants were responded to the questionnaire respectively.

### Patients group

The majority of patients were young adults, aged 20-40 years, male, and having a history of COVID-19 contact/close contact and in-home isolation (Table 1).

**Table 1:**
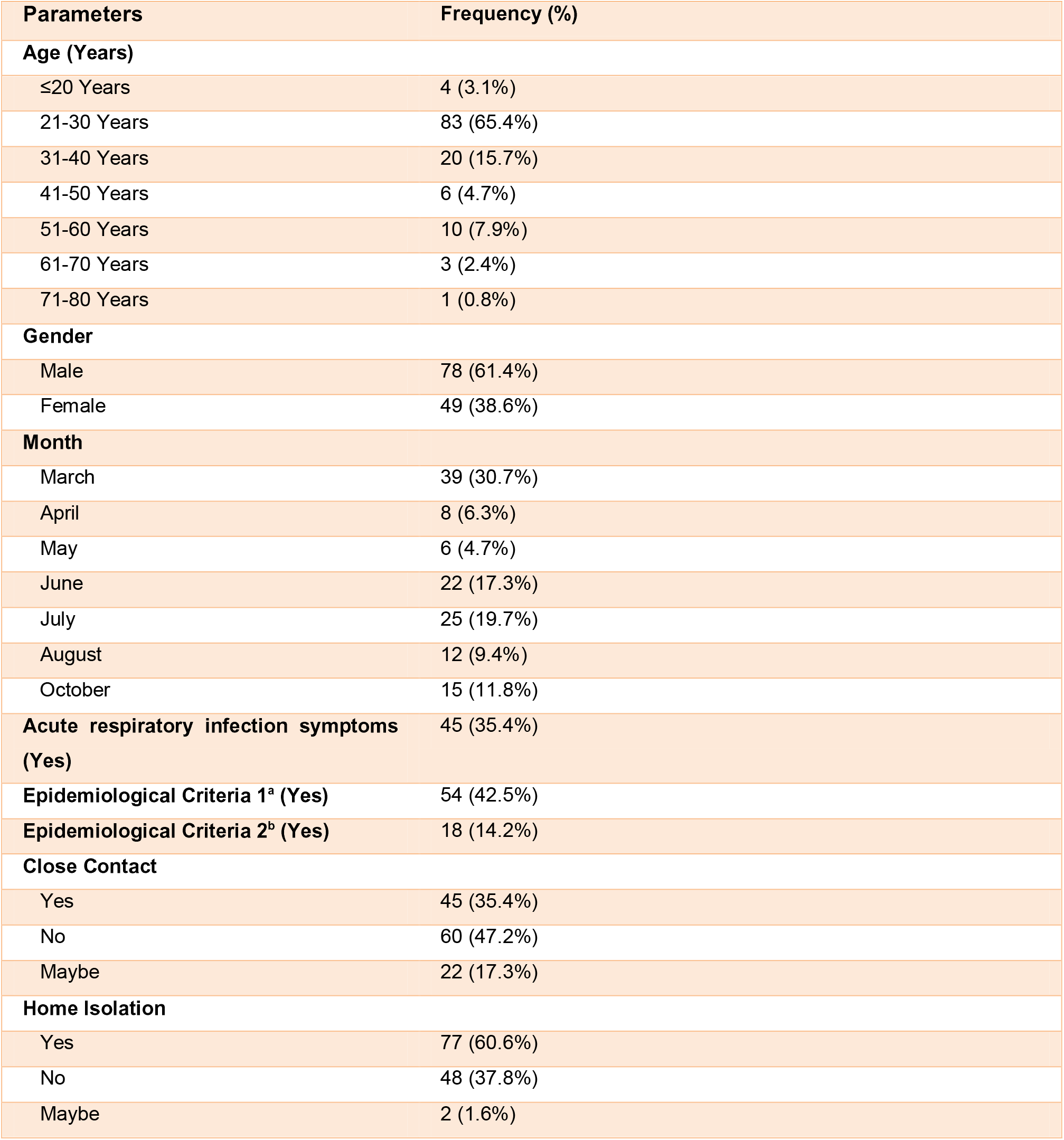
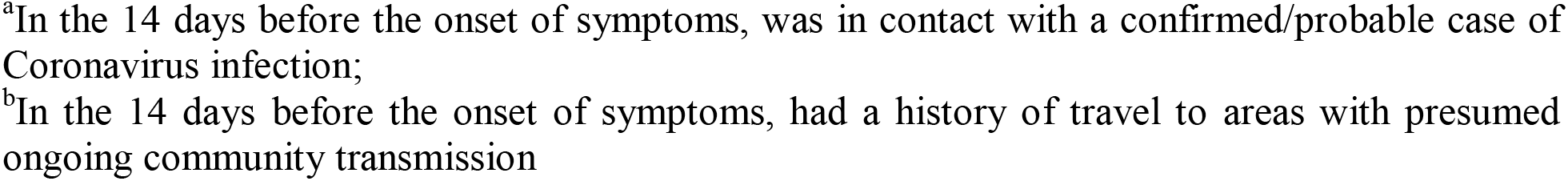
Descriptive characteristics of COVID-19 patient population (suspects/confirmed/contacts)

|  |  |
| --- | --- |
| **Parameters** | **Frequency (%)** |
| **Age (Years)** |  |
| ≤20 Years | 4 (3.1%) |
| 21-30 Years | 83 (65.4%) |
| 31-40 Years | 20 (15.7%) |
| 41-50 Years | 6 (4.7%) |
| 51-60 Years | 10 (7.9%) |
| 61-70 Years | 3 (2.4%) |
| 71-80 Years | 1 (0.8%) |
| **Gender** |  |
| Male | 78 (61.4%) |
| Female | 49 (38.6%) |
| **Month** |  |
| March | 39 (30.7%) |
| April | 8 (6.3%) |
| May | 6 (4.7%) |
| June | 22 (17.3%) |
| July | 25 (19.7%) |
| August | 12 (9.4%) |
| October | 15 (11.8%) |
| **Acute respiratory infection symptoms (Yes)** | 45 (35.4%) |
| **Epidemiological Criteria 1<sup>a</sup> (Yes)** | 54 (42.5%) |
| **Epidemiological Criteria 2<sup>b</sup> (Yes)** | 18 (14.2%) |
| **Close Contact** |  |
| Yes | 45 (35.4%) |
| No | 60 (47.2%) |
| Maybe | 22 (17.3%) |
| **Home Isolation** |  |
| Yes | 77 (60.6%) |
| No | 48 (37.8%) |
| Maybe | 2 (1.6%) |

<sup>a</sup>In the 14 days before the onset of symptoms, was in contact with a confirmed/probable case of Coronavirus infection;
<sup>b</sup>In the 14 days before the onset of symptoms, had a history of travel to areas with presumed ongoing community transmission

### FEAR

Individuals with no travel history had a greater degree of fear (Fig. 1A). A certain level of fear of contracting the disease from HCWs was also prevalent in the general population. Fear of death due to COVID-19 infection was significantly greater in the age group of 51-60 years whereas those in the 41–50-year age group had a higher incidence of bad dreams and nightmares.

**Fig 1A:**
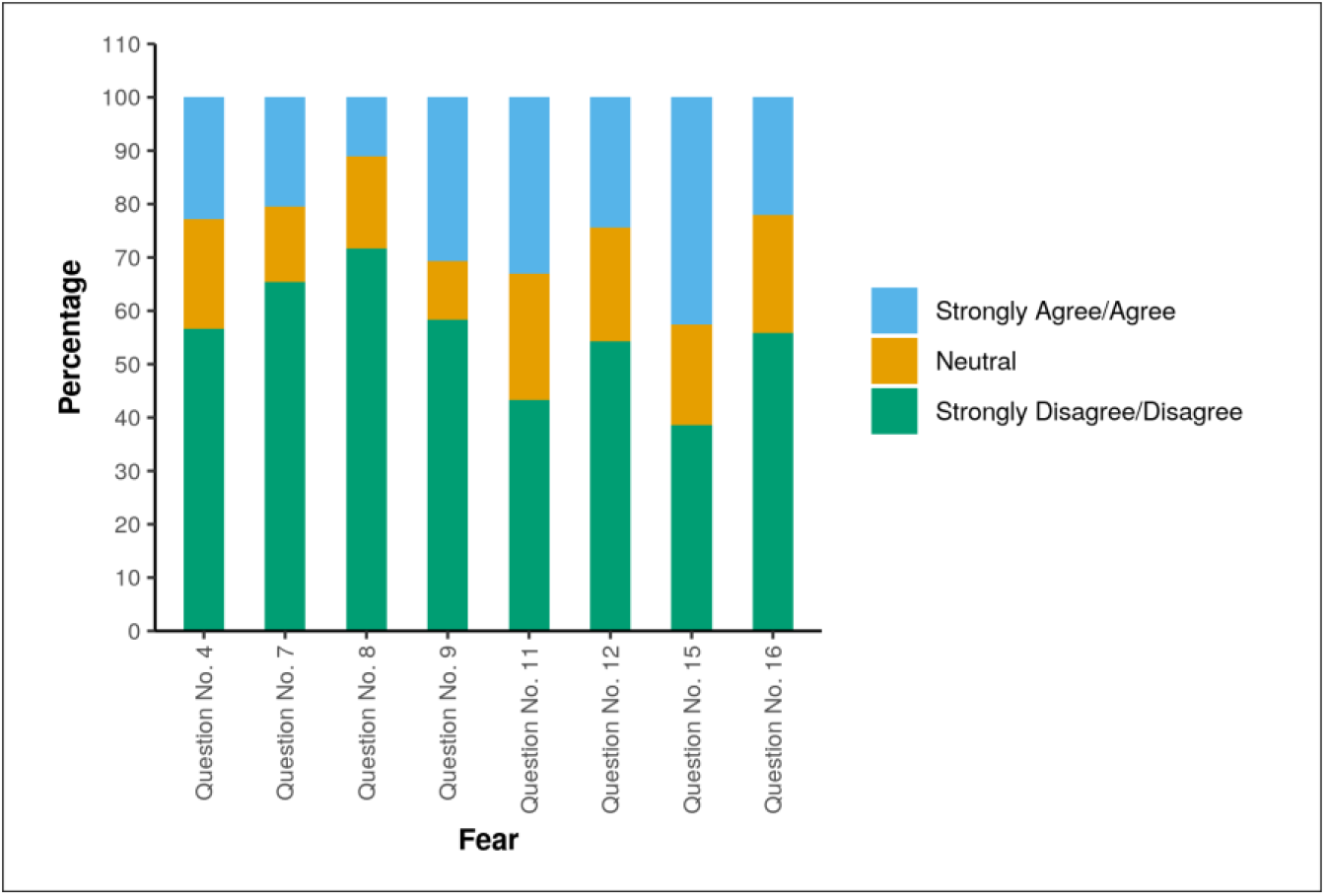
Multibar diagram representing the response to questions assessing fear in a patient population.

### ANXIETY

Higher levels of anxiety were observed in symptomatic subjects in the form of difficulty breathing in the absence of any physical exertion (Fig. 2A). The age group 41-50 were found to have significant anxiety with difficulty in relaxing and close to panicking. Home isolated/quarantined individuals had higher levels of anxiety manifesting as awareness of dryness of mouth and difficulty in breathing in the absence of physical exertion.

**Fig 2A:**
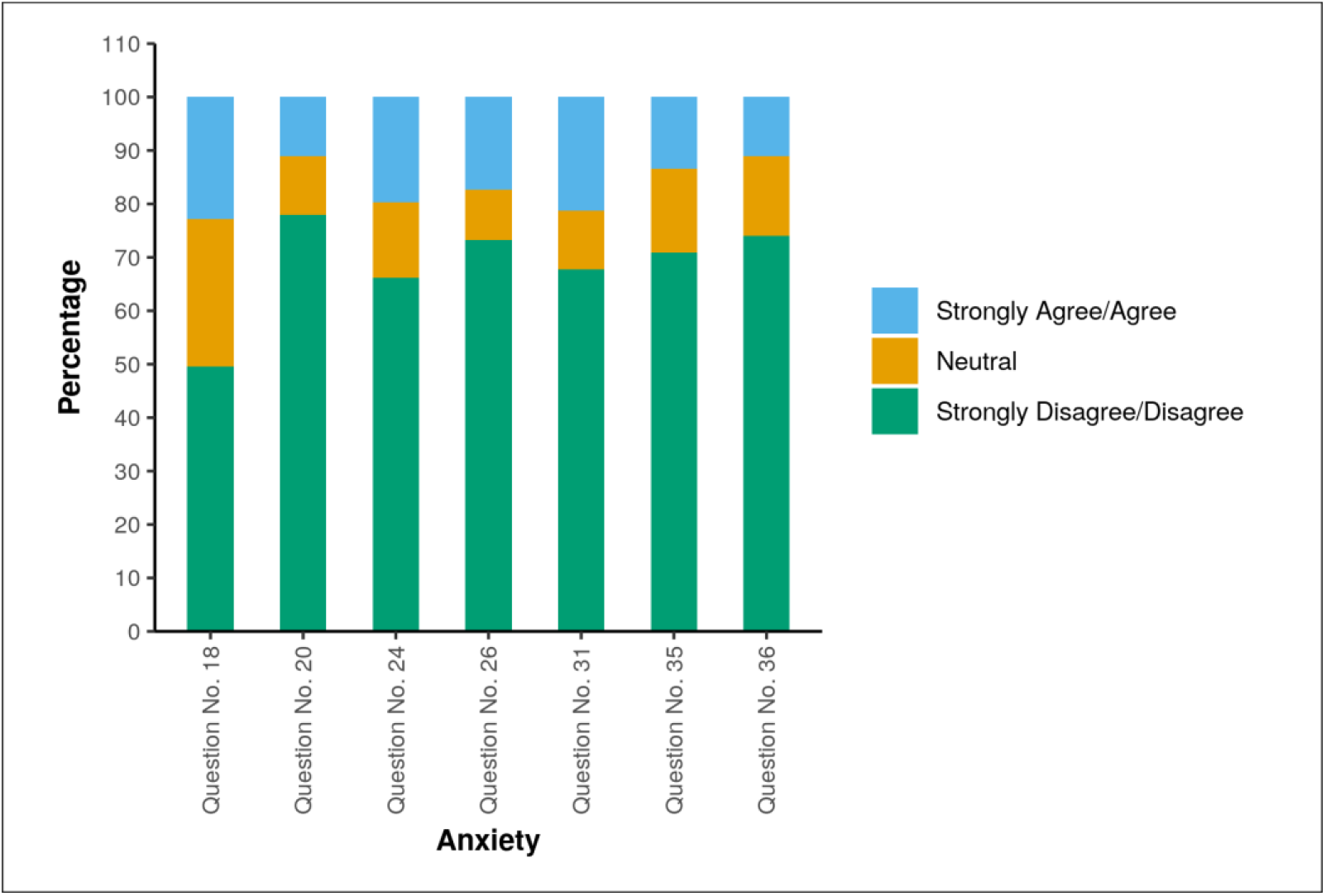
Multibar diagram representing the response to questions assessing anxiety in a patient population.

### STRESS

Greater stress levels were observed among subjects who were home isolated or quarantined juxtaposed to those who were not (Fig. 3A). Participants with acute respiratory symptoms reported stress in the form of trembling. Subjects in the age group 41-50 predominantly complained of feeling nervous, worried in certain situations of panic, tendency to overreact, inability to control things in life, inability to overcome difficulties in daily life, and coping with things. Higher levels of stress were noted in subjects with a history of travel to areas with ongoing community transmission. These individuals reported an inability to control important things in life, as well as the stress of unemployment during or after the disease. Those who visited the hospital reported stress in the form of trembling. The month of June noticed a high proportion of subjects who reported being upset due to things happening unexpectedly, inability to overcome difficulties in daily life, and anger at things being outside their control. A significant number of individuals complained of agitation in March as the lockdown phase began.

**Fig 3A:**
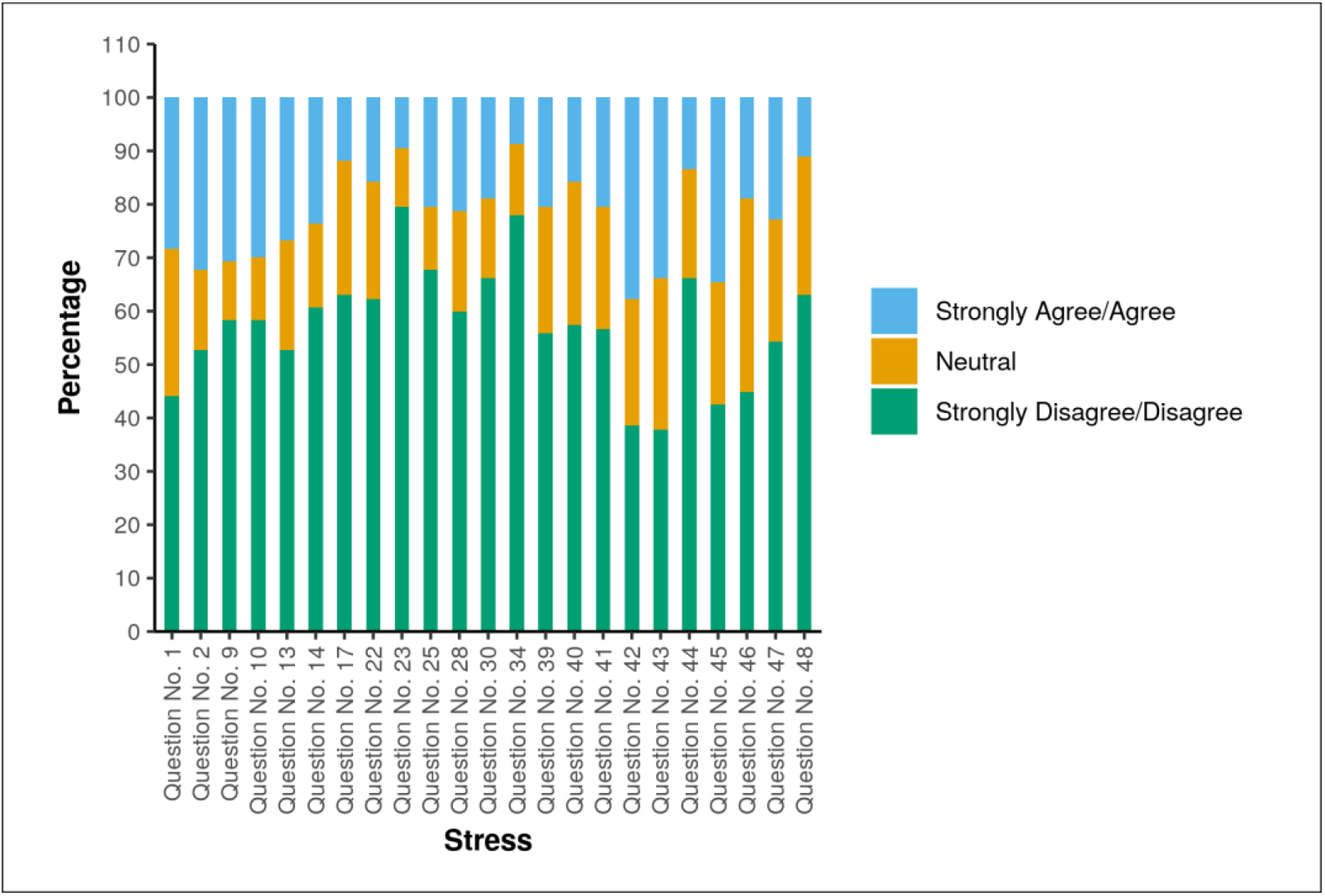
Multibar diagram representing the response to questions assessing stress in a patient population.

### DEPRESSION

Participants with symptoms of acute respiratory infection reported worthlessness (Fig. 4A). Those between the ages of and 41-50 complained of a feeling of nothing to look forward to, and a lack of enthusiasm. Subjects with a history of close contact with a positive case reported feeling downhearted and blue. A lack of initiative to do things was noted among subjects who were home isolated or quarantined. The month of June witnessed the highest number of subjects feeling worthless and nothing to look forward to, while participants in May had a significantly higher incidence of lack of initiative to do things.

**Fig 4A:**
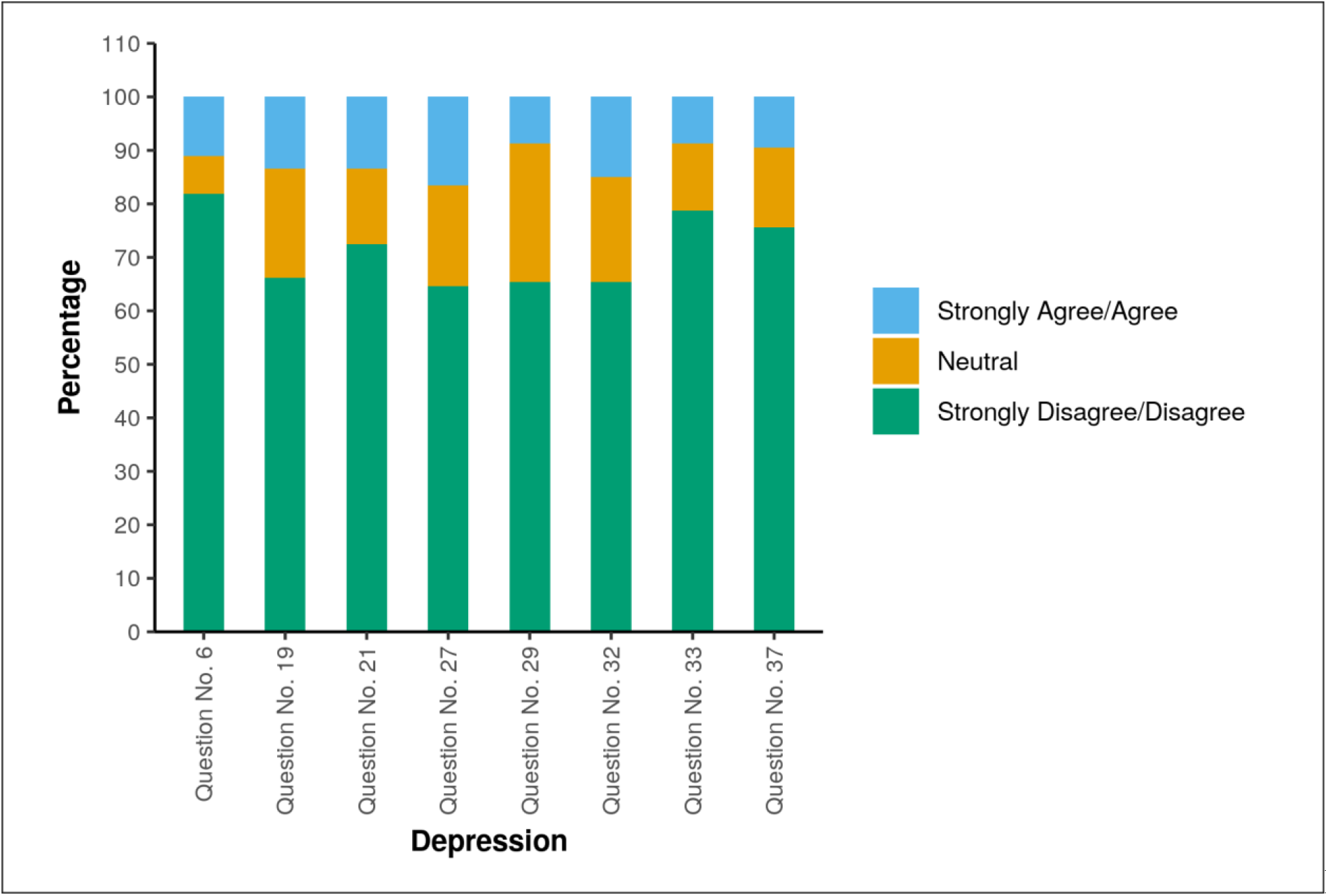
Multibar diagram representing the response to questions assessing depression in a patient population.

No patients group were followed up due to lack of response.

### Healthcare workers group

The majority of HCWs were young adults, aged 20-40 years, male, resident workers, working in non-COVID area, and following all COVID-19 precautions (Table 2).

**Table 2:** Descriptive characteristics of health care worker population.

|  |  |
| --- | --- |
| Parameters | Frequency (%) |
| **Age (Years)** |  |
| 21-30 | 141 (78.3%) |
| 31-40 | 36 (20.0%) |
| 41-50 | 3 (1.7%) |
| **Gender** |  |
| Male | 119 (66.1%) |
| Female | 61 (33.9%) |
| **Designation** |  |
| Resident | 149 (82.8%) |
| Nurse | 18 (10.0%) |
| Professor | 11 (6.1%) |
| Fellows | 1 (0.6%) |
| PhD Scholar | 1 (0.6%) |
| **Area of Work** |  |
| COVID | 54 (30.0%) |
| NON-COVID | 126 (70.0%) |
| **Caring for COVID Confirmed cases** |  |
| Yes | 28 (15.6%) |
| No | 136 (75.6%) |
| Maybe | 16 (8.9%) |
| **Following All Precautions** |  |
| Yes | 139 (77.2%) |
| No | 35 (19.4%) |
| Maybe | 6 (3.3%) |
| **Acute respiratory infection Symptoms After Contact** |  |
| Yes | 8 (4.4%) |
| No | 165 (91.7%) |
| Maybe | 7 (3.9%) |
| **Close Contact with COVID-19 Positive Case (Yes)** | 39 (21.7%) |
| **History of Travel (Yes)** | 8 (4.4%) |

### FEAR

The fear of contracting COVID-19 infection was observed in 18.3% of subjects, while 51.7% of HCWs reported fear of transmitting the infection to their family members and loved ones (Fig. 1B). No statistically significant difference was noted among the various groups in terms of fear during the pandemic.

**Fig 1B:**
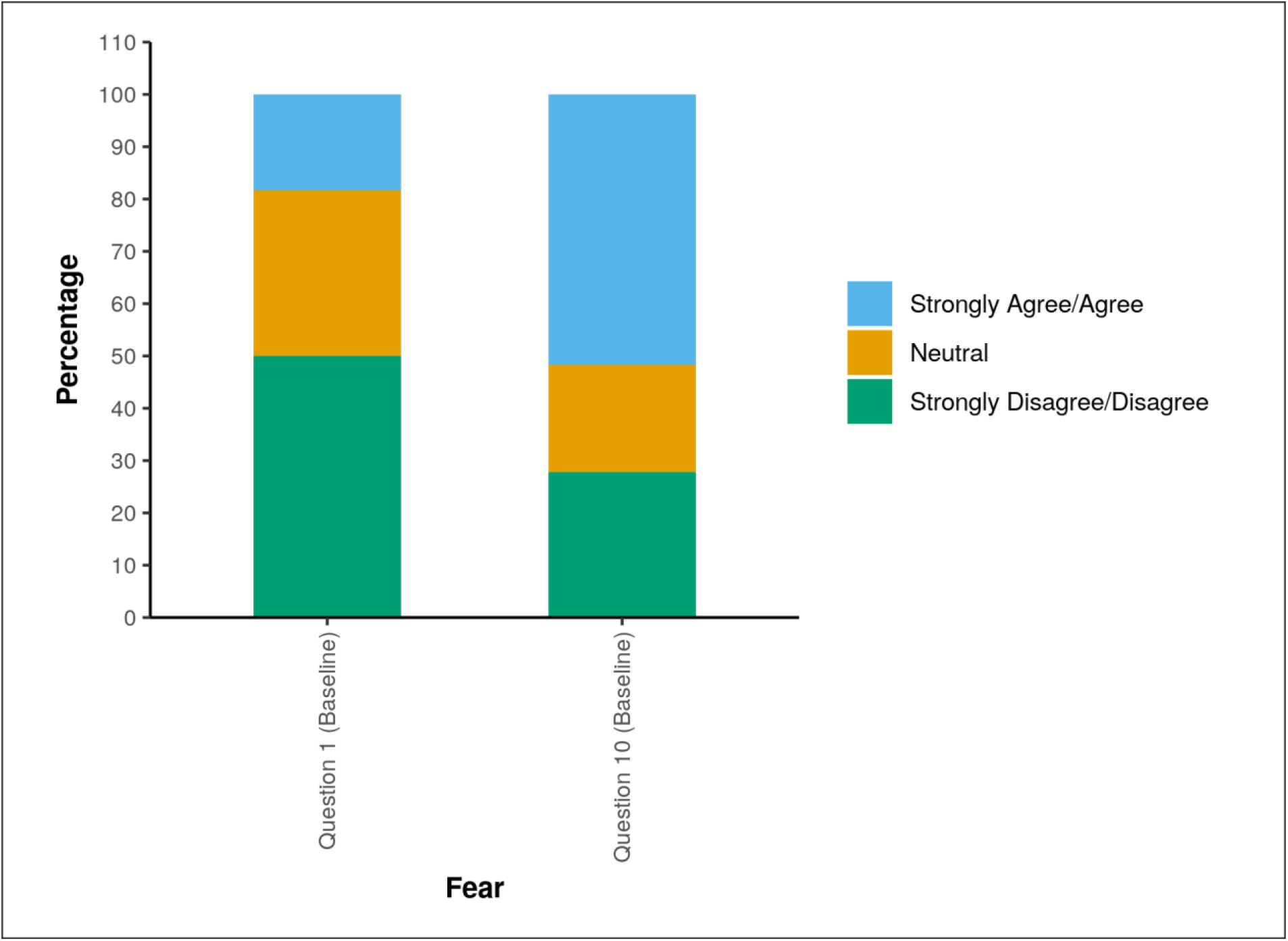
Multibar diagram representing the response to questions assessing fear in HCW population.

### ANXIETY

A significant number of subjects had a history of acute respiratory symptoms, and 14.4% of subjects reported dryness of the mouth (Fig. 2B). Palpitation, tremors, feeling scared without any reason, and tendency to panic was reported by HCWs who were working in designated COVID wards. HCWs with a history of contact with a COVID positive/suspect case claimed that they experienced dryness of their mouth, felt scared without any reason, and were close to panic.

**Fig 2B:**
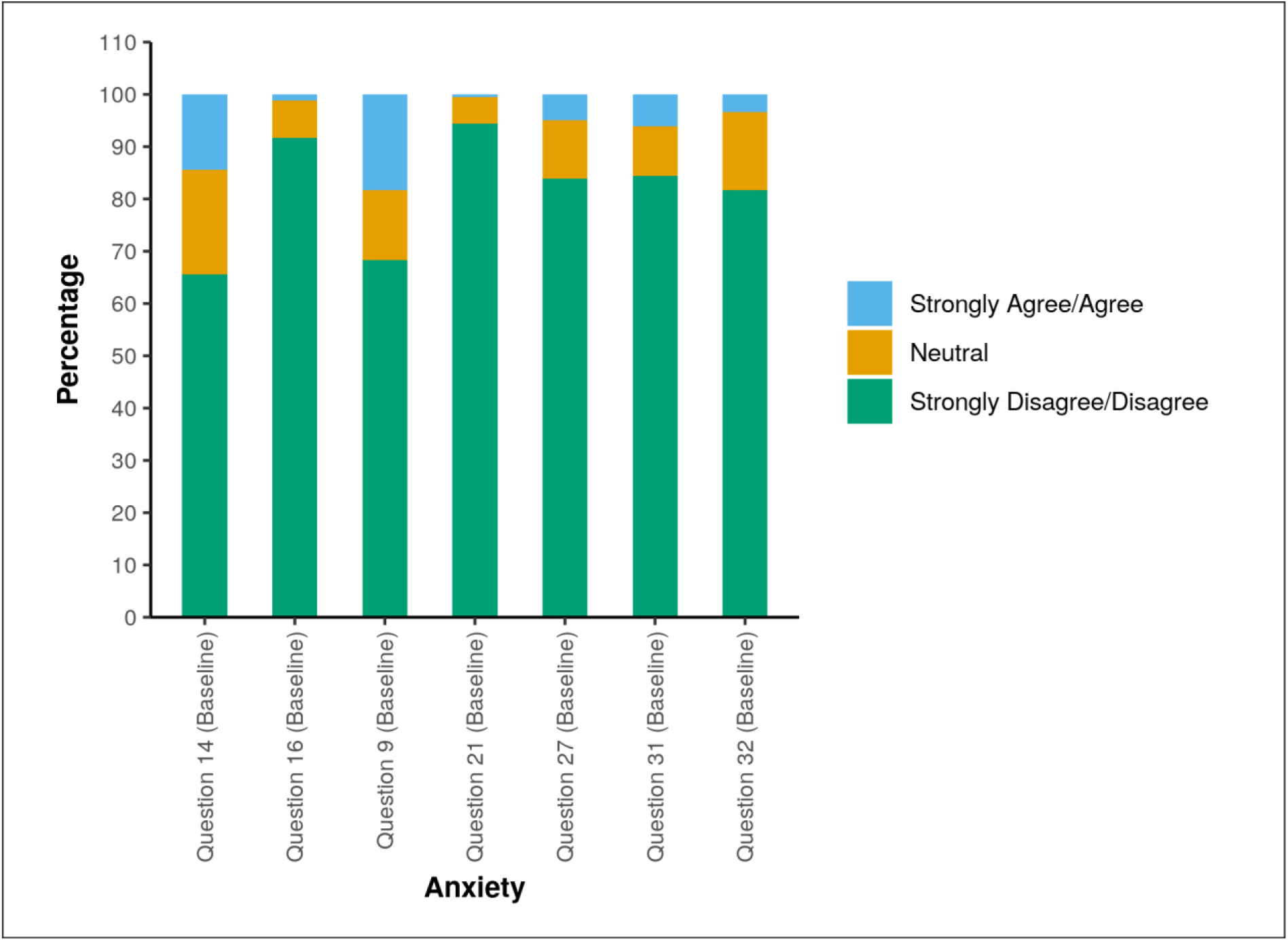
Multibar diagram representing the response to questions assessing anxiety in HCW population.

### STRESS

HCWs with acute respiratory symptoms reported significant incapability to cope with things and were unable to control their irritability (Fig. 3B). HCWs working in COVID-19 positive/suspect wards felt they were using a lot of nervous energy and found themselves getting agitated. Among the various categories of HCWs, nurses had the highest incidence of feeling rather touchy, while professors had the least. Residents reported feeling a lot of anger when things were out of control. Some positivity and optimism were noted among HCWs who felt they were following all necessary precautions and they felt things were going their way during the pandemic, while HCWs who reported not following precautions found it difficult to control their irritabilities in life. Those who were not sure if they were taking steps for prevention often felt angered about things being out of their control. Female HCWs had a higher proportion of feeling hard to relax juxtaposed to their male counterparts. HCWs with a history of contact with a positive case had a higher incidence of feeling agitated, touchy, nervous, and stressed, and communicated with difficulty in trying to wind down. Among those with a history of contact, higher levels of stress were observed among HCWs who developed upper respiratory tract symptoms after exposure.

**Fig 3B:**
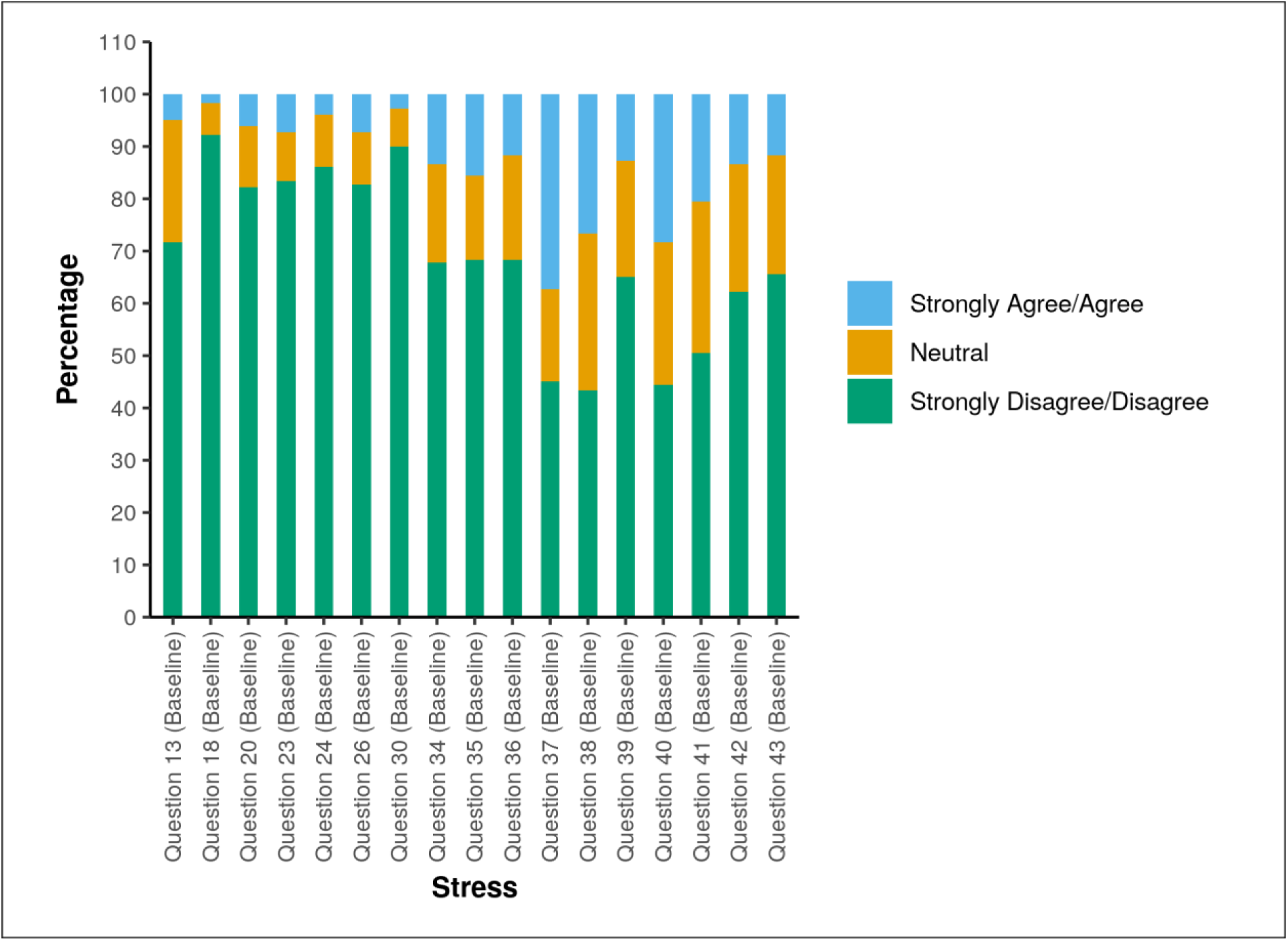
Multibar diagram representing the response to questions assessing stress in HCW population.

### DEPRESSION

A higher tendency of depression was observed in HCWs posted in COVID-19-positive/suspect areas or with history contact with a positive/suspected case, as they described feeling downhearted and blue with a lack of enthusiasm and having nothing to look forward to (Fig. 4B). Of interest, those with no history of exposure to positive cases had a higher proportion of being unable to experience any positive feeling at all. A greater incidence of lack of enthusiasm was prevalent among male HCWs compared to their female counterparts.

**Fig 4B:**
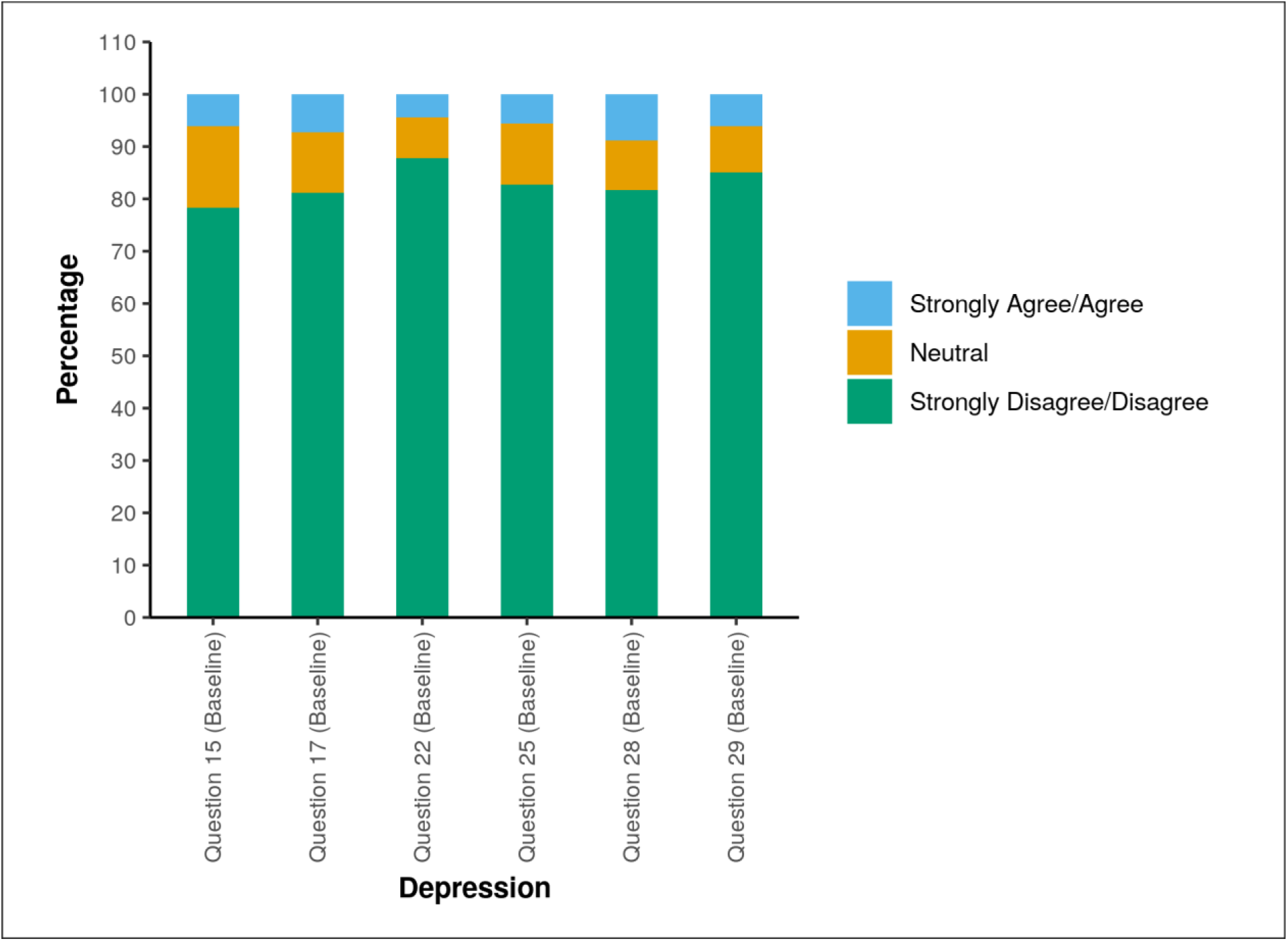
Multibar diagram representing the response to questions assessing depression in HCW population.

### Healthcare Workers (follow-up)

Although a large number of HCWs denied feeling depressed as a result of social abandonment and had less trouble relaxing than at the time of the first contact, no statistically significant variations in reaction to other psychological health criteria were observed.

## Discussion

In our research, we discovered that fear was found to be higher among people who had no previous travel background, and the general public was afraid of catching the disease from healthcare workers and vice versa. The fear of death was found to be higher in the 51-60 age group, while nightmares were found to be higher in the 41-50 age group.

HCWs with respiratory symptoms or those working in the COVID-19 ward found it particularly difficult to wind down, which caused great uncertainty and stigmatization for both staff and patients (12). 18.3% of people were afraid of developing COVID-19 infection. Around half of the HCWs said they were afraid of spreading the infection to their families and loved ones. Anxiety was higher among those who were quarantined at home, with the most common symptoms being shortness of breath, even when not exerted, and dry mouth. The isolated and home quarantine populations had higher stress levels, which manifested as a proclivity to overreact, failure to regulate emotions, nervousness, and trouble dealing with everyday life. Nurses were the most stressed, while professors were the least stressed. Females HCWs had a harder coping time than their male counterparts. Male HCWs were found to have a higher rate of lack of enthusiasm than their female counterparts. HCWs who worked in COVID-19 designated wards reported higher levels of stress, depression, and anxiety juxtaposed to their counterparts. HCWs who had previous contact with a COVID positive/suspected case reported dry mouth, feeling frightened for no apparent cause, and being on the verge of panic.

The symptoms of posttraumatic stress disorder (PTSD) and depression were observed in 28.9% and 31.2% of respondents, respectively. In a previous study conducted during the SARS pandemic, longer periods of quarantine were linked to an increased incidence of PTSD symptoms (13). In contrast, in our study, a history of travel or unemployment was found to be substantially correlated with these factors. The agitation parameter was found to be highest in March and June 2020, that is, in the early months of the pandemic. The depression parameter was highest in June and was linked to feelings of worthlessness and blueness among the respondents, in contrast to a study where higher levels of depression were during the first few weeks of the outbreak and the initial lockdown period i.e. March-April (14). These results can also be linked to post-SARS 2003 outbreak findings, which showed that among SARS survivors, the prevalence of post-recovery likely or clinician-diagnosed anxiety disorder, depressive disorder, and post-traumatic stress disorder (PTSD) was 19%, 20%, and 28%, respectively (15). These findings are in contrast with those of a similar report on HCWs and the psychological effects of the SARS 2003 pandemic, which discovered that high-risk HCWs had elevated stress levels. Despite their trust in infection-control measures, more high-risk HCWs recorded exhaustion, poor sleep, concern about health, and fear of social interaction, which were not significantly different from levels in low-risk HCWs control subjects (16).

A series of previous studies regarding the mental health impact of infectious disease pandemics and outbreaks have been conducted, with the majority being during the SARS and influenza pandemics of 2003 and 2009, respectively. One of these found a large range in prevalence rates of mental health problems, such as anxiety, depression, post-traumatic stress symptoms, or disorders, might be especially prominent among HCWs and survivors that are directly affected by epidemics and face a real threat of infection and difficult circumstances such as isolation/quarantine or difficult working conditions (18). Another study illustrated that a novel deadly infectious disease, such as SARS, can cause significant prolonged psychiatric problems (19). It has also been found that during the outbreak, SARS survivors among HCWs had stress levels similar to those of non-HCWs; however, HCWs showed significantly higher stress levels in 2004 and had higher depression, anxiety, posttraumatic symptoms, and GHQ-12 scores (20). A similar study conducted again during the SARS pandemic found that HCWs reported significantly more positive and more negative psychological effects from SARS than did control subjects. HCWs declared confidence in infection-control measures (21). Looking at the patients’ side, another previous study found that general stress and negative psychological effects are increased in SARS patients, particularly among infected HCWs (22).

Efficient preparation and support, the creation of material and emotional reserves, effective leadership, and measures to ensure the well-being of HCWs and their families will be significant motivators to come to work (23-24). Before SARS or another virulent disease breaks out, it is important to think about how to better help HCWs through timely knowledge exchange, proper infection prevention protocols, income protection during outbreaks, risk management for family members, minimizing disruptions to other clinical services, and the degree of psychosocial distress. In the case of a COVID-19 (SARS-CoV 2) pandemic, this could help to reduce staff absenteeism and, as a result, hospital services would be less disrupted. They suggest that psychological services could be important in the rehabilitation phase and should not be forgotten, as we face the evolving new outbreak of COVID-19 virus. Our results highlight the need to enhance HCWs’ preparedness and competence in detecting and managing the psychological sequelae of future comparable infectious disease outbreaks. It has also shown higher levels of PTSD-like symptoms in both the HCWs and COVID-19 survivor populations are prevalent. The risk factor analysis can not only improve the detection of undiagnosed psychiatric complications but also provide insight into a possible model of care delivery for COVID -19 survivors.

This study is limited by the collection of data over a long duration of time, and the varying trends of the pandemic over the months may have caused a variation in psychological perception among the participants. Also, there was attrition and many participants were lost to follow-up. The use of self-reported questionnaires for fear and anxiety aspects may also be considered a drawback in comparison to clinician-diagnosed psychological sequelae. However, the results provide important insights for policymakers and healthcare professionals, involved in clinical work as well as hospital administration, around the world while delineating the psychological sequelae of life-threatening and fatal infectious disease outbreaks. It is imperative to understand that a pandemic is bound to have long-term psychiatric sequelae and our work doesn’t end with the subsidence of the pandemic. The psychological sequelae may appear further with a delay as delayed complications of covid-19 are yet to be recognized and understood.

## Conclusion

Our study suggests that all the parameters ranging from fear, anxiety, stress, and depression were significantly high compared to those experienced during the SARS 2003 outbreak, thus emphasizing the psychological aspects of the COVID-19 pandemic. These results emphasize a need for strong preparedness and readiness to counter the ongoing as well as any future pandemics on mental beings. Our results also suggest significant psychological impact to both patient as well as HCW population; thus warranting adequate prevention measures and ample psychological support to these population groups. There is an immense amount of further research and studies required in these domains in the future to obtain complete facts around the pivotal aspects of the mental impact of any pandemics in history as well as in the future.

## Data Availability

It is available with the corresponding author, once required, will be provided.

## Contributors

AP, KV, HK, N contributed to the data collection, analysis, and was involved in manuscript writing. VSR, YAB, DK, PKP gave the concept, interpreted analysis, critically reviewed the draft, and approved for publication along with all authors.

## Data sharing

It will be made available to others as required by requesting to corresponding author.

## Acknoweldgement

None

## References

1. Li Q, Guan X, Wu P, Wang X, Zhou L, Tong Y, et al. Early Transmission Dynamics in Wuhan, China, of Novel Coronavirus–Infected Pneumonia. N Engl J Med. 2020 Mar 26;382(13):1199–207.

2. World Health Organization. 2020. Statement on the Second Meeting of the International Health Regulations (2005) Emergency Committee Regarding the Outbreak of Novel Coronavirus (2019-nCoV) Published January 30

3. Bao Y, Sun Y, Meng S, Shi J, Lu L. 2019-nCoV epidemic: address mental health care to empower society. The Lancet. 2020 Feb;395(10224):e37–8.

4. Ni MY, Yang L, Leung CMC, Li N, Yao XI, Wang Y, et al. Mental Health, Risk Factors, and Social Media Use During the COVID-19 Epidemic and Cordon Sanitaire Among the Community and Health Professionals in Wuhan, China: Cross-Sectional Survey. JMIR Ment Health. 2020 May 12;7(5):e19009.

5. Bo H-X, Li W, Yang Y, Wang Y, Zhang Q, Cheung T, et al. Posttraumatic stress symptoms and attitude toward crisis mental health services among clinically stable patients with COVID-19 in China. Psychol Med. 2020 Mar 27;1–2.

6. Nguyen HC, Nguyen MH, Do BN, Tran CQ, Nguyen TTP, Pham KM, et al. People with Suspected COVID-19 Symptoms Were More Likely Depressed and Had Lower Health-Related Quality of Life: The Potential Benefit of Health Literacy. J Clin Med. 2020 Mar 31;9(4):965.

7. Cai H, Tu B, Ma J, Chen L, Fu L, Jiang Y, et al. Psychological impacts and coping strategies of front-line medical staff during COVID-19 outbreak in Hunan, China. Med Sci Monit [Internet]. 2020 Mar 23 [cited 2021 May 21];26. Available from: https://www.medscimonit.com/abstract/index/idArt/924171

8. Lai J, Ma S, Wang Y, Cai Z, Hu J, Wei N, et al. Factors Associated With Mental Health Outcomes Among Health Care Workers Exposed to Coronavirus Disease 2019. JAMA Netw Open. 2020 Mar 23;3(3):e203976.

9. Zhang W, Wang K, Yin L, Zhao W, Xue Q, Peng M, et al. Mental Health and Psychosocial Problems of Medical Health Workers during the COVID-19 Epidemic in China. Psychother Psychosom. 2020;89(4):242–50.

10. Cohen S, Kamarck T, Mermelstein R. A Global Measure of Perceived Stress. J Health Soc Behav. 1983 Dec;24(4):385.

11. Lovibond, S. H., & Lovibond, P. F. (1995). Manual for the Depression Anxiety Stress Scales (2nd ed.). Sydney: Psychology Foundation of Australia.

12. Maunder R, Hunter J, Vincent L, Bennett J, Peladeau N, Leszcz M, et al. The immediate psychological and occupational impact of the 2003 SARS outbreak in a teaching hospital. CMAJ. 2003 May 13;168(10):1245–51.

13. Hawryluck L, Gold WL, Robinson S, Pogorski S, Galea S, Styra R. SARS Control and Psychological Effects of Quarantine, Toronto, Canada. Emerg Infect Dis. 2004;10(7):7

14. Lee SM, Kang WS, Cho A-R, Kim T, Park JK. Psychological impact of the 2015 MERS outbreak on hospital workers and quarantined hemodialysis patients. Compr Psychiatry. 2018 Nov;87:123–7.

15. Chau SWH, Wong OWH, Ramakrishnan R, Chan SSM, Wong EKY, Li PYT, et al. History for some or lesson for all? A systematic review and meta-analysis on the immediate and long-term mental health impact of the 2002–2003 Severe Acute Respiratory Syndrome (SARS) outbreak. BMC Public Health. 2021 Dec;21(1):670.

16. McAlonan GM, Lee AM, Cheung V, Cheung C, Tsang KW, Sham PC, et al. Immediate and Sustained Psychological Impact of an Emerging Infectious Disease Outbreak on Health Care Workers. Can J Psychiatry. 2007 Apr;52(4):241–7.

17. Tsang HW, Scudds RJ, Chan EY. Psychosocial impact of SARS. Emerg Infect Dis. 2004;10(7):1326–1327. doi:10.3201/eid1007.040090

18. Zürcher SJ, Kerksieck P, Adamus C, Burr CM, Lehmann AI, Huber FK, et al. Prevalence of Mental Health Problems During Virus Epidemics in the General Public, Health Care Workers and Survivors: A Rapid Review of the Evidence. Front Public Health. 2020 Nov 11;8:560389.

19. Mak IWC, Chu CM, Pan PC, Yiu MGC, Chan VL. Long-term psychiatric morbidities among SARS survivors. Gen Hosp Psychiatry. 2009 Jul;31(4):318–26.

20. Lee AM, Wong JG, McAlonan GM, Cheung V, Cheung C, Sham PC, et al. Stress and Psychological Distress among SARS Survivors 1 Year after the Outbreak. Can J Psychiatry. 2007 Apr;52(4):233–40.

21. Chua SE, Cheung V, Cheung C, McAlonan GM, Wong JW, Cheung EP, et al. Psychological Effects of the SARS Outbreak in Hong Kong on High-Risk Health Care Workers. Can J Psychiatry. 2004 Jun;49(6):391–3.

22. Chua SE, Cheung V, McAlonan GM, Cheung C, Wong JW, Cheung EP, et al. Stress and Psychological Impact on SARS Patients during the Outbreak. Can J Psychiatry. 2004 Jun;49(6):385–90.

23. Maunder RG, Leszcz M, Savage D, Adam MA, Peladeau N, Romano D, et al. Applying the Lessons of SARS to Pandemic Influenza: An Evidence-based Approach to Mitigating the Stress Experienced by Healthcare Workers. Can J Public Health. 2008 Nov;99(6):486–8.

24. Psychological First Aid: Field Operations Guide: 2nd Edition: (536202011-001) [Internet]. American Psychological Association; 2006 [cited 2021 May 21]. Available from: http://doi.apa.org/get-pe-doi.cfm?doi=10.1037/e536202011-001

