## Supplemental Appendix 1-2 for "Fear, Anxiety, Stress, and Depression of novel coronavirus (COVID-19) pandemic among patients and their healthcare workers"

**APPENDIX 1: Questionnaire for patient populations**

| NO | QUESTION |
| --- | --- |
| 1 | Are/Were you experiencing any Stress due to isolation? |
| 2 | Are/Were you experiencing any Stress before results of the tests for nCov 19? |
| 3 | In the last two weeks, how often have you felt difficulties were piling up so high that you could not overcome them? |
| 4 | Are/Were you experiencing any Stress due to social abandonment/humiliation? |
| 5 | Are you having any Lack of sleep or sleep problems recently? |
| 6 | Have you ever thought of harming yourself in some way recently? |
| 7 | Are you having any Fear of dying due to nCov infection? |
| 8 | Are having any bad dreams or nightmares recently? |
| 9 | Are you facing any trouble in relaxing or restlessness recently? |
| 10 | Do you have any Stress of contracting disease from family/ friends/ strangers? |
| 11 | Do you have any Fear of contracting disease from any Health Care Worker in the hospital? |
| 12 | Do you have any Fear of contracting other hospital acquired infections from your stay in hospital? |
| 13 | Do you have any Stress of expenditure in hospital during your stay? |
| 14 | Do you have any Stress of being unemployed during OR after disease? |
| 15 | Do you have any Fear of infecting the loved ones/relatives/people in contact with you? |
| 16 | Do you have any Fear of not having a healthy future OR any residual impact of disease? |
| 17 | I found it hard to wind down |
| 18 | I was aware of dryness of mouth |
| 19 | I couldn’t seem to experience any positive feeling at all |
| 20 | I experienced difficulty breathing [excessively rapid breathing, breathlessness in absence of any physical exertion] |
| 21 | I found it difficult to work up the initiative to do things |
| 22 | I tended to overreact to situations |
| 23 | I experienced trembling |
| 24 | I felt that I was using a lot of nervous energy |
| 25 | I was worried about situations in which I might panic and make a fool of myself |
| 26 | I felt I had nothing to look forward to |
| 27 | I found it difficult to relax |
| 28 | I found myself getting agitated |
| 29 | I felt downhearted and blue |
| 30 | I was intolerant of anything that kept me from getting on with what I was doing |
| 31 | I felt I was close to panic |
| 32 | I was unable to become enthusiastic about anything |
| 33 | I felt I wasn’t worth much as a person |
| 34 | I felt that I was rather touchy |
| 35 | I was aware of the action of my heart in the absence of physical exertion [sense of heart rate increase, heart missing a beat] |
| 36 | I felt scared without any good reason |
| 37 | I felt that life was meaninglessly to |
| 38 | Any other problems faced Stress |
| 39 | In the last two weeks, how often have you been upset because of something that happened unexpectedly? |
| 40 | In the last two weeks, how often have you felt that you were unable to control important things in life? |
| 41 | In the last two weeks, how often did you feel nervous and stressed? |
| 42 | In the last two weeks, how often have you felt confident about your ability to handle your personal problems? |
| 43 | In the last two weeks, how often have you felt things were going your way? |
| 44 | In the last two weeks, how often have you found that you could not cope with all the things you had to do? |
| 45 | In the last two weeks, how often have you been able to control irritations in your life? |
| 46 | In the last two weeks, how often have you felt that you were on top of things? |
| 47 | In the last two weeks, how often have you been angered because of things that were outside of your control? |
| 48 | In the last two weeks, how often have you felt difficulties were piling up so high that you could not overcome them? |

**APPENDIX 2: Questionnaire for healthcare workers**

| NO. | QUESTION |
| --- | --- |
| 1 | Are/Were you experiencing any Stress due to isolation? |
| 2 | Are/Were you experiencing any Stress before results of the tests for nCov 19? |
| 3 | In the last two weeks, how often have you felt difficulties were piling up so high that you could not overcome them? |
| 4 | Are/Were you experiencing any Stress due to social abandonment/humiliation? |
| 5 | Are you having any Lack of sleep or sleep problems recently? |
| 6 | Have you ever thought of harming yourself in some way recently? |
| 7 | Are you having any Fear of dying due to nCov infection? |
| 8 | Are having any bad dreams or nightmares recently? |
| 9 | Are you facing any trouble in relaxing or restlessness recently? |
| 10 | Do you have any Stress of contracting disease from family/ friends/ strangers? |
| 11 | Do you have any Fear of contracting disease from any Health Care Worker in the hospital? |
| 12 | Do you have any Fear of contracting other hospital acquired infections from your stay in hospital? |
| 13 | Do you have any Stress of expenditure in hospital during your stay? |
| 14 | Do you have any Stress of being unemployed during OR after disease? |
| 15 | Do you have any Fear of infecting the loved ones/relatives/people in contact with you? |
| 16 | Do you have any Fear of not having a healthy future OR any residual impact of disease? |
| 17 | I found it hard to wind down |
| 18 | I was aware of dryness of mouth |
| 19 | I couldn’t seem to experience any positive feeling at all |
| 20 | I experienced difficulty breathing [excessively rapid breathing, breathlessness in absence of any physical exertion] |
| 21 | I found it difficult to work up the initiative to do things |
| 22 | I tended to overreact to situations |
| 23 | I experienced trembling |
| 24 | I felt that I was using a lot of nervous energy |
| 25 | I was worried about situations in which I might panic and make a fool of myself |
| 26 | I felt I had nothing to look forward to |
| 27 | I found it difficult to relax |
| 28 | I found myself getting agitated |
| 29 | I felt downhearted and blue |
| 30 | I was intolerant of anything that kept me from getting on with what I was doing |
| 31 | I felt I was close to panic |
| 32 | I was unable to become enthusiastic about anything |
| 33 | I felt I wasn’t worth much as a person |
| 34 | I felt that I was rather touchy |
| 35 | I was aware of the action of my heart in the absence of physical exertion [sense of heart rate increase, heart missing a beat] |
| 36 | I felt scared without any good reason |
| 37 | I felt that life was meaninglessly to |
| 38 | Any other problems faced Stress |
| 39 | In the last two weeks, how often have you been upset because of something that happened unexpectedly? |
| 40 | In the last two weeks, how often have you felt that you were unable to control important things in life? |
| 41 | In the last two weeks, how often did you feel nervous and stressed? |
| 42 | In the last two weeks, how often have you felt confident about your ability to handle your personal problems? |
| 43 | In the last two weeks, how often have you felt things were going your way? |
| 44 | In the last two weeks, how often have you found that you could not cope with all the things you had to do? |
| 45 | In the last two weeks, how often have you been able to control irritations in your life? |
| 46 | In the last two weeks, how often have you felt that you were on top of things? |
| 47 | In the last two weeks, how often have you been angered because of things that were outside of your control? |
| 48 | In the last two weeks, how often have you felt difficulties were piling up so high that you could not overcome them? |
